# When ownership is not enough: Evaluating the co-development and uptake of the Options Assessment Toolkit (OAT) for *Plasmodium vivax* radical cure in malaria-endemic countries

**DOI:** 10.64898/2026.05.06.26352599

**Authors:** Bijaya Shrestha, Neena Valecha, Kamala Thriemer, Caroline Anita Lynch

## Abstract

**Introduction:** Radical cure of *Plasmodium vivax* malaria remains a major challenge in endemic countries. New treatment options add opportunities but also increase complexity of policy decisions. Malaria treatment policies are often shaped by World Health Organisation (WHO) guidance, limiting scope for local adaptation. The Options Assessment Toolkit (OAT) was developed to support national policy making for the radical cure of vivax malaria. This study evaluates its co-development, uptake and practical influence.

**Methods:** We conducted a qualitative study with stakeholders in Afghanistan, the Solomon Islands, and Vietnam (co-developers) and Nepal (new user context). In-depth interviews were conducted with representatives from the National Malaria Programs between January and March 2025. Thematic analysis was conducted using NVivo 12, guided by pre-defined research questions, and reported according to COREQ criteria.

**Results:** Participants described a strong ownership resulting from the inclusive co-development process. However, none of the co-developing countries used the OAT for formal policy decision-making. Instead, it primarily supported structured deliberation, planning, and contextual appraisal of emerging treatment options. A key finding was the conflation between decision-support and implementation feasibility: the toolkit’s perceived value was often judged according to whether recommended strategies could be operationalised within existing system constraints. Instrumental uptake was shaped by institutional structures, system readiness, variation in national analytic capacity, and reliance on WHO endorsement.

**Conclusions:** The findings suggest that ownership alone is insufficient for institutional uptake. Decision-support tools exert influence primarily by reshaping deliberation, but require embedding within formal policy, regulatory, and financing processes for sustained impact.

## Introduction

Globally, malaria treatment policy and strategic approach decisions are often shaped by centralised, evidence-based guidance from the World Health Organisation’s (WHO) Global Malaria Programme (GMP), with many national programmes relying heavily on these recommendations as both a technical and political source of legitimacy (1, 2). Donor funding and technical assistance are frequently aligned with these global standards. While this centralised model has supported standardisation of treatment policies, it has also reinforced dependence on external normative authority, particularly in settings with limited domestic evidence appraisal capacity (2).

We previously developed the Options Assessment Toolkit (OAT) as a decision-support instrument in response to the first major expansion of tools for *Plasmodium vivax* malaria in more than six decades (3–5). Since then, the global malaria landscape has shifted further, with renewed commitments to elimination by 2030 and a growing pipeline of new drugs, diagnostics, and delivery approaches. At the same time, the WHO has moved toward a “living guidance” model, resulting in more frequent updates to technical recommendations (6). Together, these developments have increased the number and complexity of decisions facing National Malaria Control Programs (NMCPs), particularly in relation to case management and chemoprevention strategies (7–9). These demands coincide with declining global malaria financing and reduced availability of multilateral technical advice which has reduced technical support at global, regional, and country levels to help programmes interpret guidance and integrate emerging evidence into policy and practice (10).

In response to the prioritisation by NMCPs for better support for policy makers in the Asia Pacific the OAT was co-developed between 2020 and 2022 as a structured decision-support framework for *P. vivax* radical cure (4). The OAT was intended to help countries choose new treatments and diagnostics depending on local conditions and health system readiness, and support translation of anticipated global guidelines into national policy (11). At the time of development there was no global guidance on the use of tafenoquine or high-dose primaquine, but their integration in future recommendations was anticipated. The OAT was designed as a facilitative framework to support deliberation, identify implementation gaps and enable anticipatory planning.

Limited evidence exists on how the OAT was used or integrated into national malaria policy processes following its development (3). More broadly, there is little empirical evidence on how decision-support tools function within hierarchical global health governance systems where policy change is mediated by regulatory pathways, implementation feasibility, external donor views and reliance on WHO endorsement. This study assesses the experience of co-development and early use of the OAT in Afghanistan, the Solomon Islands, and Vietnam and assesses its application in Nepal, which did not participate in co-development. Furthermore, this study examines how ownership, institutional structures, and external legitimacy mechanisms shaped the uptake and influence of the OAT.

## Materials and methods

### Design, population and sample

This study employed a qualitative approach involving the original co-developing countries; Afghanistan, the Solomon Islands and Vietnam (Table S1) to explore their experience of the OAT development process. Participants in these countries were NMCP representatives who had been directly involved in the co-development of the OAT. The evaluation therefore intentionally followed up with the same institutional actors to assess their experiences of the development process and subsequent use of the tool.

In addition, we included participants from Nepal to explore how the tool was perceived and used outside the original co-developing settings. This allowed comparison between co-development-driven ownership and independent engagement with the toolkit.

Given the institutional focus of the study, participants were purposively sampled as individuals occupying senior technical or managerial roles within the NMCPs and directly involved in malaria policy formulation or advisory processes. The aim was not numerical representativeness but to capture perspectives from actors positioned within national decision-making structures.

The study follows a standard qualitative methodology outlined by the consolidated criteria for reporting qualitative research (COREQ) checklist (Table S2) (12).

### Data collection

The principal investigators (KT and CAL) introduced BS to the co-developing NMCPs via email and BS conducted data collection between January to March 2025. A total of three virtual interviews (Microsoft Teams) were conducted with one participant from Vietnam, and four interviews each with participants from Afghanistan and the Solomon Islands. One in person interview was conducted with two respondents from Nepal.

Virtual interviews were conducted in English. In the first meeting, BS introduced himself and shared his personal details and concluded by outlining the objectives of forthcoming meetings. These initial meetings lasted approximately half an hour each. Participants also shared their personal details and informal context beyond the main topic of research. From the second meeting onward, interview guidelines were used to structure the sessions, which typically lasted between 45 to 90 minutes. The interview with the participants from Nepal were conducted by BS in Nepali.

The interview guidelines were organised around several key themes. First, it aimed to explore the participants’ experiences with and ownership of the OAT. It also examined the co-development process of the OAT, along with strategies to support its uptake and integration into current policy decision making. Finally, the interview guidelines focused on understanding policy change pathways, including the main barriers and enabling factors. For Nepal, the interview included an exploratory discussion in which participants were briefed on the OAT and guided through each question in relation to their own programme context.

Data collection was concluded when no new information emerged on the pre-defined interview themes. Interviews were audio-recorded using Krisp-AI software and subsequently transcribed. The Nepali interview was directly transcribed into English. Transcripts were reviewed for accuracy and shared with the participants to allow them to provide clarifications or additional input.

### Data analysis

Following the transcription, the data were coded line by line. Analysis combined deductive coding aligned with the predefined research objectives and inductive coding to capture emergent themes beyond the original framework (Table S3) (13). Thematic analysis was conducted with attention to how participants framed institutional constraints, legitimacy, and feasibility, rather than solely describing reported use of the toolkit.

NVivo 12 (QSR International, Melbourne, Vic., Australia) was used to manage and analyze the data (14). Preliminary findings were discussed with the research team through weekly meetings to refine interpretation and enhance analytic credibility.

### Reflexivity

BS conducted the interviews and had no prior formal role in the development of the OAT, which may have reduced desirability bias in participant responses. KT and CAL were involved in the original OAT development and contributed contextual and policy expertise during interpretation. Divergent interpretations were discussed within the team to enhance analytical rigour.

### Patient and Public Involvement

Patients and members of the public were not involved in the design, conduct, reporting, or dissemination of this research. The study focused on national malaria programme stakeholders as institutional decision-makers rather than patient-level perspectives.

### Ethical approval

The protocol outlining the OAT developed was reviewed and approved by the Human Research Ethics Committee of NT Health and Menzies School of Health Research in 2022, and an amendment for the evaluation was received in 2024 (2022-4245). Verbal consent was obtained from all study participants.

## Results

The results are presented according to key themes that emerged from the analysis: 1) OAT ownership from the co-development process; 2) conceptual use versus instrumental uptake; 3) conflation between decision-support and implementation; 4) reliance on WHO as a legitimacy mechanism.

### OAT ownership from the co-development process

Participants from the Solomon Islands and Afghanistan described strong ownership of the OAT arising from the co-development process. Although each participant represented their country individually, the regular engagement with other country representatives and the research team helped sustained momentum without becoming burdensome.

Participants emphasised that the OAT did not feel preconceived and that their input was reflected in the evolving tool. The incorporation of feedback reinforced perceptions that the tool was responsive to national contexts rather than externally imposed.

> *“The OAT development process was very good and consultative because at each step, we were involved, and the documents were shared with us.”*
>
> — -NMP, Afghanistan

Participants also valued the facilitation approach. The process was described as highly interactive, with participants noting that face-to-face engagement might have further strengthened collaboration, but that virtual facilitation remained effective.

> *“I think I would rate it as a 9 (out of 10). The whole process of development would have been 10, if it was in-person. Sometimes the experience of discussing online is different from discussing in-person.”*
>
> — -NMP, Solomon Islands

### Conceptual use versus instrumental uptake

The OAT was originally intended to guide evidence-based decision-making, particularly in the context of revising radical cure strategies for *P. vivax* malaria. Despite strong ownership, none of the co-developing countries reported using the OAT to directly revise national malaria treatment policy. Rather than being formally adopted as a policy instrument, the OAT was primarily used as a thinking tool that supported internal dialogue, anticipatory planning, and reflections on system readiness. In Afghanistan and the Solomon Islands, participants noted that the co-development process itself influenced their thinking and informed discussions on programmatic gaps, diagnostics, and treatment strategies.

> *“I think I was more excited that such a tool was being developed. For me, it made the role of a manager easier when thinking about strategies for anything in this case; Vivax.”*
>
> — -NMP, Solomon Islands

Participants described the OAT as useful for structuring conversations about change and evidence uptake, even when formal policy action remained constrained. In Nepal, where the OAT had not been previously introduced, it supported dialogue on data gaps, procedural steps, and system bottlenecks relevant to future policy change.

> *“The OAT helps us understand what needs to change, but following through with approvals requires time and persistence.”*
>
> — - NMP, Nepal

Furthermore the OAT was also used as a checklist or planning aid, particularly to identify operational bottlenecks and prioritise tasks. Participant noted that this was particularly relevant where basic health system components such as diagnostics, trained personnel or pharmacovigilance were weak,.

> *“Without a basic system in place, the OAT became more of a checklist than a roadmap.”*
>
> — -NMP, Afghanistan

Beyond functional use, perceptions of OAT’s added value varied by context and user experience. A participant from Vietnam shared that the national malaria guideline was referenced during development of OAT, which contributed to the OAT’s clarity and ease of application.

> *“The OAT, I don’t think it is much different from what we have in each country.”*
>
> — -NMP, Vietnam

### Conflation between decision-support and implementation

A central finding was that participants frequently evaluated the OAT not as a decision-support framework in isolation, but in relation to whether its recommended treatment options could be implemented within existing system constraints. Participants frequently evaluated the toolkit’s usefulness based on whether its recommended options could be implemented within existing system constraints.

> *“It’s not the tool, it’s what it recommends, and how that fits into our system.”*
>
> — -NMP, Afghanistan; echoed in Nepal

Where implementation of high dose primaquine or tafenoquine was perceived as infeasible; due to diagnostic gaps particularly the absence of G6PD testing, human resource capacity, supply chain procurement challenges, weak pharmacovigilance and follow-up systems, enthusiasm for engaging with the OAT diminished. These concerns were not about the OAT itself, but about whether the health system was ready to act on what the OAT suggested.

> *“To successfully use the OAT, we need more than just administrative capacity; we also need trained staff at health facilities, diagnostic tools like G6PD kits, and financial support to cover associated costs.”*
>
> — -NMP, Afghanistan

Supply chain constraints further limited feasibility of implementing recommended treatment options, particularly because tafenoquine was difficult to procure in the small quantities required by national programmes.

> *“Logistics make this harder because companies are unwilling to supply small quantities.”*
>
> — -NMP, Vietnam

Concerns about patient safety and risk aversion were closely linked to limitations in system capacity, including weak pharmacovigilance, inadequate follow-up mechanisms, and constrained referral pathways. In such contexts, stakeholders expressed hesitation to advance new treatment options without confidence that adverse events could be promptly detected, managed, and communicated.

> *“Patient safety and cases (mortality) is our main concern. Therefore, we are cautious to implement new strategy immediately.”*
>
> — -NMP, Solomon Islands

### Reliance on WHO as a legitimacy mechanism

Across settings, reliance on WHO guidance strongly shaped perceptions of when and how policy change could occur. Participants described WHO endorsement as a prerequisite for advancing treatment revisions, particularly in contexts with limited internal capacity for independent evidence appraisal.

> *“We follow WHO recommendations because they go through rigorous review. Our country doesn’t have the capacity to challenge or re-interpret those recommendations.”*
>
> — -NMP, Solomon Islands

WHO recommendations were viewed not only as technical guidance but as a source of institutional protection. Endorsement reduced perceived political and professional risk associated with introducing new treatment strategies, particularly those involving safety considerations such as G6PD testing or shorter radical cure regimens. Participants were therefore reluctant to advance OAT-recommended options independently. Reliance on WHO recommendations remained central, particularly in settings with limited internal evidence review capacity.

However, in Nepal participants described a different perspective. While WHO guidance remained central, participants described the OAT as supporting contextual interpretation rather than replacing global guidance.

> *“WHO recommendations are global but our context is local.”*
>
> — -NMP, Nepal

> *“Earlier, we always waited for WHO protocol and followed it strictly. But this can encourage us to think for ourselves and became a ‘thinking tool’.”*
>
> — -NMP, Nepal

## Discussion

This study demonstrates that co-development and perceived ownership do not automatically translate into institutional uptake of decision-support tools. Although the OAT was regarded as contextually relevant, it was not directly used for policy revision. Its instrumental uptake was limited by system readiness, regulatory processes, and reliance on WHO endorsement. However, the toolkit facilitated structured reflection on treatment options, system readiness, and future policy directions, even in settings where immediate guideline revision was not feasible. Our findings suggest that the effectiveness of decision-support tools depends less on participatory design alone and more on how they are embedded within governance structures.

Current national strategic planning in many malaria programmes follows a structured cycle, typically involving a malaria program or mid-term programme review followed by revision of the National Strategic Plan (NSP). These reviews are largely retrospective, assessing performance against existing targets and indicators (15). They do not systematically incorporate horizon scanning for emerging tools, evolving evidence, or innovations not yet formally endorsed within country, but for which the country could prepare. Structured engagement platforms may partially address this gap. NMCPs cannot realistically participate in all global research generating relevant evidence, particularly as innovation pipelines expand. Mechanisms that create space for collective reflection on emerging data, discussion of contextual transferability, and appraisal of system implications may serve as catalysts for proactive thinking (16). Even when not resulting in immediate policy revision, such processes may strengthen preparedness and institutional confidence in navigating evolving guidance.

Participants consistently valued the OAT’s iterative co-development process. Structured facilitation, visible incorporation of feedback, and repeated cycles of deliberation fostered legitimacy and a sense of ownership. Participatory design has been shown to enhance acceptability, trust and perceived relevance of policy instruments (17–20). Importantly the value of sstructured engagement extended beyond ownership. In this study, the OAT enabled systematic reflection on implementation gaps and emerging therapeutic options, even where immediate policy change was not feasible. This form of influence reflects conceptual models of knowledge utilization, where impact occurs through reframing, agenda-setting, and capacity strengthening rather than immediate policy change (21–23).

A central implication of this study is that ownership alone is insufficient for operationalisation. While participants felt ownership over the OAT, its utilisation in practice was limited. Co-development improves perceived relevance but did not, on its own, overcome systemic constraints on implementation (17, 24, 25). Policy tools operate within political and institutional structures that shape their uptake. Formal adoption often depends on regulatory pathways, approval hierarchies, budget cycles, and risk tolerance (22, 26). These structural factors are closely linked to variation in national capacity to independently appraise and synthesize evidence. In Vietnam, established technical structures and guideline processes reduced the perceived added value of the OAT. In contrast, other settings expressed greater reliance on external guidance for evidence interpretation (2). This variability suggests that decision-support tools cannot be uniformly designed or implemented, but must account for differing levels of analytic capacity, regulatory maturity, and institutional autonomy. Evaluating decision-support tools solely on immediate policy shifts may therefore underestimate their impact (27, 28). In complex health systems, incremental shifts in how evidence is interpreted and discussed likely represent meaningful progress. However, for sustained influence, such tools must likely be embedded within formal policy and planning cycles, including mid-term programme reviews, NSP revisions, and associated budgeting and implementation processes.

The conflation between analytic appraisal and implementation mandate emerged as an important barrier to instrumental uptake of the OAT. In several settings, the OAT was perceived as implicitly endorsing specific treatment changes rather than as a structured appraisal framework. This is a broader pattern in global health governance, where technical guidance is frequently interpreted as prescriptive within hierarchical, compliance-oriented systems (22, 29, 30). When implementation conditions were not in place, engagement shifted toward problem framing rather than progression to adoption. This form of conceptual use is well described in the policy literature, where influence often occurs through problem framing and agenda setting and reframing rather than direct regulatory change (31, 32).

Progression towards operational use of the OAT was limited primarily by the feasibility of implementing the treatment options identified. Countries were reluctant to progress toward radical cure policy changes when foundational system elements, particularly G6PD testing, were not yet in place. Similar feasibility constraints have been identified as key determinants of policy adoption for malaria diagnostics and treatment innovations (24, 33). Decision-support tools may therefore benefit from explicitly integrating readiness mapping and phased implementation pathways. In settings with limited domestic investment and reliance on external financing (34, 35), operational constraints may outweigh analytic appraisal. An important question emerging from this work is whether structured, nationally anchored deliberation may strengthen long-term political commitment and domestic financing. While this study cannot empirically test that proposition, literature on policy ownership and fiscal commitment suggests that domestically anchored decisions may enhance accountability and financing incentives (22).

WHO endorsement functioned as both epistemic authority and political risk mitigation. In contexts with limited domestic capacity for evidence appraisal, WHO recommendations provided credibility and reduced perceived liability associated with introducing new treatment strategies (36–40). The OAT did not act in opposition to WHO guidance but complemented it by supporting contextual interpretation. The broader architecture of global health governance, in which normative authority and financing mechanisms are closely aligned, reflects this. In many settings, alignment with WHO recommendations is closely linked to funding processes, including those supported by the Global Fund (41). As a result, progression toward policy change often depends not only on analytic appraisal, but on external endorsement.

However, the global health financing environment is changing (10, 42). Budget reductions affecting WHO and financial constraints within the Global Fund ecosystem may reduce the extent of centralised normative and technical support available to countries. If external technical guidance becomes less readily accessible, reliance on endorsement as the primary pathway for policy validation may become increasingly difficult to sustain. Embedding anticipatory appraisal mechanisms within national systems, potentially through structured tools such as the OAT, could enhance resilience of malaria governance in a more constrained global architecture.

This study had several limitations. First, not all participants involved in the validation phase were the same as those in the co-development phase due to travel and health constraints, which may have created discontinuities in feedback. However, participation across both phases remained within NMCP structures. Second, this work was conducted in four countries with distinct governance and epidemiological contexts. While this diversity enriches the analysis, it may limit the transferability of findings to other institutional settings. Third, the analysis relied on self-reported perceptions, which may be influenced by recall bias or social desirability. Nevertheless, participants articulated both supportive and critical perspectives, suggesting that a range of views was captured. Finally, the study focused on a malaria-specific policy pathway. Although several governance dynamics identified here are likely relevant beyond malaria, caution is warranted in extrapolating findings to other disease areas without further empirical validation.

## Conclusion

This study demonstrates that co-development and ownership of decision support tools do not automatically translate into operational use. In malaria governance systems characterised by strong reliance on WHO endorsement and constrained implementation capacity, decision-support tools may exert influence primarily by reshaping deliberation rather than directly changing guidelines. Findings suggest that future policy tools should be explicitly designed for dual function: supporting anticipatory appraisal while integrating readiness mapping and financing pathways.

## Data Availability

Relevant anonymized data are included in the manuscript. Due to sensitive nature of patient information, raw data such as transcripts and notes cannot be shared publicly. However, data may be made available upon reasonable request by contacting

## Acknowledgement

The research team acknowledges the participants of the research

## Supporting information

Table S1: Country context

Table S2: Consolidated criteria for reporting qualitative research (COREQ) checklist

Table S3: Codebook for analysis

